# Baseline functional connectivity predicts who will benefit from neuromodulation: evidence from primary progressive aphasia

**DOI:** 10.1101/2024.04.19.24305354

**Authors:** Zeyi Wang, Jessica Gallegos, Donna Tippett, Chiadi U Onyike, John E Desmond, Argye E Hillis, Constantine E Frangakis, Brian Caffo, Kyrana Tsapkini

**Affiliations:** Department of Biostatistics, Johns Hopkins School of Public Health, Baltimore, MD, USA; Department of Neurology, Johns Hopkins Medicine, Baltimore, MD, USA; Department of Physical Medicine & Rehabilitation, Johns Hopkins Medicine, Baltimore, MD, USA; Department of Psychiatry and Behavioral Sciences, Johns Hopkins Medicine, Baltimore, MD, USA; Department of Cognitive Science, Johns Hopkins Medicine, Baltimore, MD, USA; Neuroscience Program, Johns Hopkins University, Baltimore, MD, USA; Department of Radiology, Johns Hopkins Medicine, Baltimore, MD, USA

**Keywords:** prediction, heterogeneity, primary progressive aphasia, transcranial direct current stimulation, inferior frontal gyrus, semantic retrieval, verbal fluency, semantic fluency, precision medicine

## Abstract

**Background:** Identifying the characteristics of individuals who demonstrate response to an intervention allows us to predict who is most likely to benefit from certain interventions. Prediction is challenging in rare and heterogeneous diseases, such as primary progressive aphasia (PPA), that have varying clinical manifestations. We aimed to determine the characteristics of those who will benefit most from transcranial direct current stimulation (tDCS) of the left inferior frontal gyrus (IFG) using a novel heterogeneity and group identification analysis.

**Methods:** We compared the predictive ability of demographic and clinical patient characteristics (e.g., PPA variant and disease progression, baseline language performance) vs. functional connectivity alone (from resting-state fMRI) in the same cohort.

**Results:** Functional connectivity alone had the highest predictive value for outcomes, explaining 62% and 75% of tDCS effect of variance in generalization (semantic fluency) and in the trained outcome of the clinical trial (written naming), contrasted with <15% predicted by clinical characteristics, including baseline language performance. Patients with higher baseline functional connectivity between the left IFG (opercularis and triangularis), and between the middle temporal pole and posterior superior temporal gyrus, were most likely to benefit from tDCS.

**Conclusions:** We show the importance of a baseline 7-minute functional connectivity scan in predicting tDCS outcomes, and point towards a precision medicine approach in neuromodulation studies. The study has important implications for clinical trials and practice, providing a statistical method that addresses heterogeneity in patient populations and allowing accurate prediction and enrollment of those who will most likely benefit from specific interventions.

## 1. Introduction

Precision medicine allows for specialized care, the minimization of unnecessary procedures, and a streamlined approach to treatment. Precision medicine implementation is emerging in language therapy interventions in aphasia (for both post-stroke and primary progressive aphasia), through prediction studies that show which patients will benefit from specific treatments are not yet clinically applicable. Some problems with prediction studies in aphasia include variability in aphasia profiles, the lack of area/symptom correspondence, the extent of damage / atrophy, and premorbid cognitive/language abilities. Addressing this heterogeneity is essential, because predicting whether and how well an individual with aphasia will respond to treatment is a key factor in developing individualized treatment plans. This need to address heterogeneity becomes even more important when considering new pharmacological, genetic, and neuromodulatory treatments that are becoming available for neurodegenerative conditions.

Our interest lies in primary progressive aphasia (PPA), a neurodegenerative disorder affecting language functions primarily [1,2], for which the only disease-modifying treatments are symptomatic language therapy and emerging neuromodulation approaches, especially transcranial direct current stimulation (tDCS) [3,4]. There is feasibility and efficacy of tDCS stimulation to trained items, as well as transfer of benefits to untrained items/words [4–6]. Investigation of factors that predict treatment response in PPA is at a nascent stage (see [7,8] for recent reviews) and is challenging, because PPA is a heterogenous clinical syndrome that includes at least three different clinical variants with varying patterns of decline [2]. Early prediction of treatment outcomes (before any treatment starts) is important in a neurodegenerative disorder because losing time means losing brain tissue.

Recent studies from different groups, including our own, have identified several baseline factors that predict efficacy of tDCS for a given patient or patient group [see [6–8] for reviews]. Five predictive factors have been identified: (1) baseline language and cognitive performance [9,10], (2) baseline cortical volume and thickness [11–13], (3) baseline white matter integrity [14], (4) clinical variant PPA [15], and (5) baseline sleep efficiency [16]. To our knowledge, no studies have utilized baseline functional brain connectivity as a predictor of tDCS effects in PPA. This is an important gap for tDCS, since evidence suggest that it modulates language outcomes in PPA is changes via functional brain connectivity changes [17–19]. Brain functional connectivity is correlated with general cognition as in Fronto-Temporal Dementia Clinical Rating Scale FTD-CDR, as we and others have reported. Finally, change in functional connectivity is one of the first alterations that happen in neurodegeneration including PPA [20,21]. Functional connectivity has been used as a predictor only in some recent tDCS studies research [22–24] that aimed to predict responses to tDCS in schizophrenia or healthy controls.

Recent studies in healthy controls highlight the specificity of functional connectivity (FC) changes after electrical stimulation [25]. We were among the first to demonstrate this particular mechanism (changes in FC) in PPA [17,19]. We found that tDCS downregulates the abnormally high FC in PPA in the areas stimulated. This hyperconnectivity was correlated with higher FTD-CDR, i.e., worse global cognition. Hyperconnectivity or hyperexcitability is a known hallmark of AD, and probably is involved in other neurodegenerative disorders [26,27]. Reductions in FC between the stimulated area (left inferior frontal gyrus, IFG) and other brain areas (such as the left middle temporal gyrus, MTG, with which it is structurally and functionally connected [28]), correlated with improvement in treatment outcomes.

Our modified outcome analysis predicts individual causal effects (e.g., had they been assigned to active tDCS vs. sham tDCS, along with oral and written naming treatment in both tDCS conditions) rather than the actual responses for the assigned intervention. Such heterogeneity analysis of tDCS in PPA patients offers an opportunity to predict the additional benefit received from active tDCS compared to sham tDCS, even before actual treatment begins. Outcomes of such analysis will be valuable to clinicians, patients, and caregivers, as it assists personalized precision health decisions.

This study closes the loop between the mechanism of tDCS and the clinical prediction of who will benefit from tDCS based on its mechanism. We hypothesized that: (1) if tDCS downregulates hyperconnectivity in stimulated areas, then baseline hyperconnectivity of the stimulated area will predict treatment outcomes. Furthermore, we hypothesized that: (2) baseline hyperconnectivity will predict tDCS effects better than other demographic or clinical factors. We tested these hypotheses using special statistical methods to account for heterogeneity in a rare disease. This study has important implications for clinical trials of potential new treatments in the spirit of precision medicine, because the methods implemented herein appear to allow us to predict the effects of these treatments before they are initiated.

## 2. Materials and Methods

### 2.1 Participants

Thirty-six patients with PPA participated in this study (17 female): 14 with logopenic variant PPA (lvPPA), 13 with non-fluent variant PPA (nfvPPA), and 9 with semantic variant PPA (svPPA). All were right-handed, native English speakers, between 50 and 80 years old, and diagnosed based on clinical assessment, neuropsychological and language testing, and MRI, according to consensus criteria [2]. Informed consent was obtained from participants or their spouses, and all data were acquired in compliance with the Johns Hopkins Hospital Institutional Review Board. Figure 1 shows recruitment and randomization to the active tDCS or sham tDCS conditions. Each PPA variant group was matched for sex, age, education, years post onset of symptoms, overall FTD-CDR score and language severity measures (Tables 1A, 1B).

**Figure 1.**
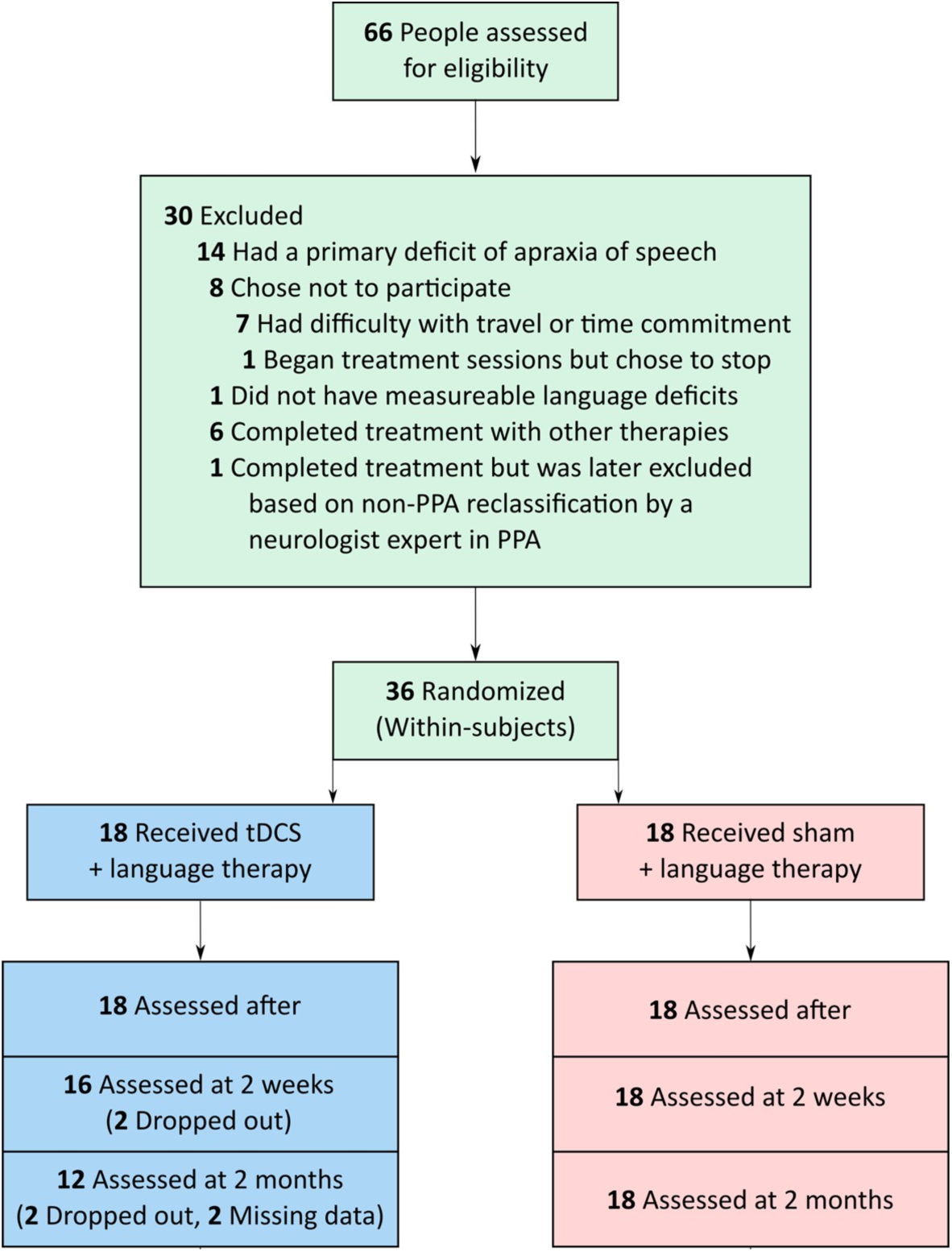
Participants recruited and randomized to active tDCS or sham tDCS.

**Table 1A.**
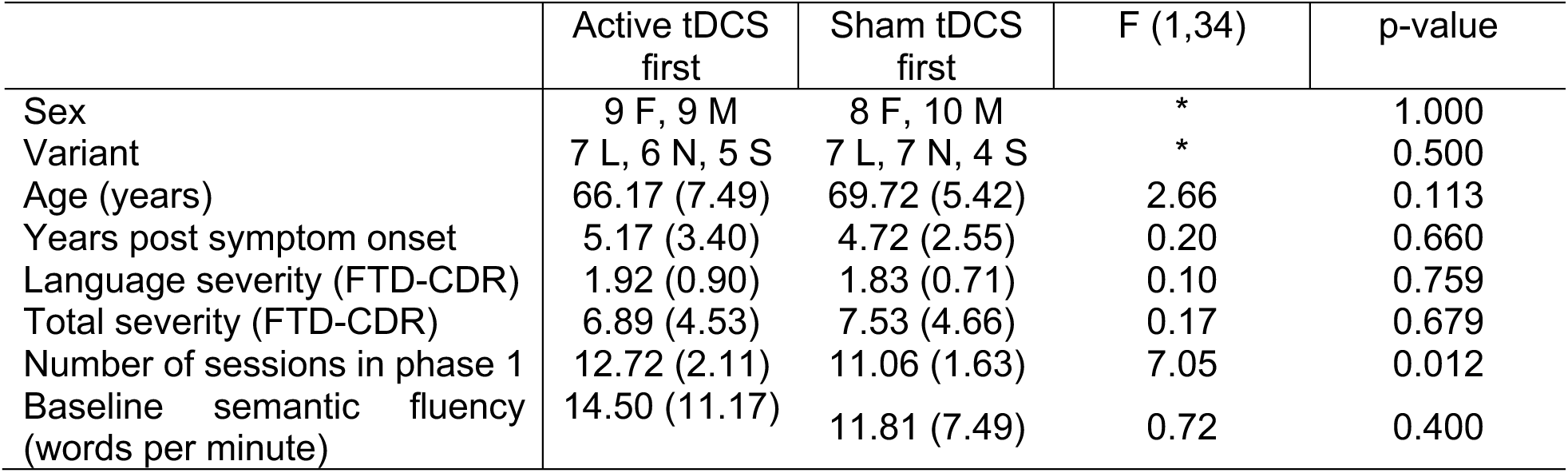
Means and standard deviations of demographic variables and baseline semantic fluency scores grouped by first-phase condition (n=36). *Fisher’s exact test used. FTD-CDR, Frontotemporal Dementia Clinical Dementia Rating Scale sum of boxes [29]. F, female; M, male. L, logopenic; N, nonfluent; S semantic.

**Table 1B.** Means and standard deviations of demographic variables and baseline semantic fluency scores grouped by PPA variant (n=36). *Fisher’s exact test used. FTD-CDR, Frontotemporal Dementia Clinical Rating Scale sum of boxes [29]. F, female; M, male. s, sham tDCS; t, active tDCS.

|  | lvPPA | nvPPA | svPPA | F(2,33) | p-value |
| --- | --- | --- | --- | --- | --- |
| Sex | 7 F, 7 M | 5 F, 8 M | 5 F, 4 M | * | 0.800 |
| First-phase condition | 7 s, 7 t | 7 s, 6 t | 4 s, 5 t | * | 1.000 |
| Age (years) | 66.29 (8.11) | 69.77 (6.00) | 67.89 (4.96) | 0.91 | 0.412 |
| Years post symptom onset | 4.82 (3.33) | 4.65 (2.66) | 5.56 (3.08) | 0.25 | 0.780 |
| Language severity (FTD-CDR) | 1.57 (0.83) | 2.04 (0.72) | 2.11 (0.78) | 1.76 | 0.188 |
| Total FTD-CDR | 6.18 (3.76) | 7.85 (4.19) | 7.89 (6.17) | 0.57 | 0.571 |
| Number of sessions in phase 1 | 11.93 (2.02) | 11.85 (1.91) | 11.89 (2.47) | 0.01 | 0.990 |
| Baseline semantic fluency (words per minute) | 17.50 (10.97) | 12.08 (8.38) | 7.94 (5.15) | 3.30 | 0.049 |

### 2.2 Overall Design

We used a within-subjects, double-blind, crossover design with two experimental conditions: speech-language therapy plus conventional anodal tDCS over the left IFG, and speech-language therapy plus sham tDCS. Each condition lasted approximately 12 consecutive weekday sessions; the two phases were separated by a 2-month wash-out period. Stratified randomization of stimulation condition within each variant determined whether each participant received active tDCS or sham tDCS first, according to our main clinical trial (ClinicalTrials.gov identifier: NCT02606422). Performance was assessed before, immediately after, two weeks after, and two months after each period of stimulation (active tDCS or sham tDCS). Participants, speech-language pathologists, and examiners were blinded to the experimental condition. In statistical analysis, we focused on first-phase data only in order to avoid impact of carryover effects due to possibly insufficient wash-out period as we had noticed when we analyzed the behavioral results[15].

### 2.3 tDCS Methods

Each daily therapy session lasted one hour. For both active tDCS and sham conditions, two 5cm x 5 cm, non-metallic, conductive, rubber electrodes covered with saline-soaked sponges were placed over the right cheek (cathodal electrode) and the left IFG centered at F7 of the EEG 10-20 electrode position (anodal electrode) [30]. The electrodes were hooked up to a Soterix 1x1 Clinical Trials device, which elicited a tingling sensation on the scalp as it ramped up within 30 seconds, to deliver current at an intensity of 2 mA (estimated current density 0.08 mA/cm^2^; estimated total charge 0.096 C/cm^2^). In the active tDCS condition, current was delivered for 20 minutes for a daily maximum of 2.4 Coulombs; in the sham tDCS condition, current ramped up to 2 mA over a 30 sec interval and immediately ramped down to elicit the same tingling sensation, a procedure that has been shown to blind participants to treatment condition [31]. Stimulation started at the beginning of each therapy session and lasted for 20 minutes; speech-language therapy continued for the full session, i.e., 25 additional minutes, for 45-50 minutes. Twice during each session, participants rated their level of pain with the Wong-Baker FACES Pain Rating Scale (www.WongBakerFACES.org).

### 2.4 Language intervention

In the present study we based our prediction methodology on generalization effects we previously found in semantic fluency [32] and not on the trained task, since trained tasks are always expected to show benefits and generalization of treatment to other tasks is the most desirable outcome in neurodegenerative disorders. Also, we have extensively reported on the results for the trained task, oral and written naming [4]. We replicated the prediction analysis for the trained task and reported the results in Appendix 5.

### 2.5 Imaging methods

Of the 36 participants, 29 had magnetic resonance imaging (MRI) scans—five were severely claustrophobic and two had pacemakers and were therefore excluded. MRI scans took place at the Kennedy Krieger Institute at Johns Hopkins University. Magnetization-prepared rapid acquisition gradient echo (MPRAGE) and resting-state functional MRI (rsfMRI) scans were acquired before treatment on a 3-Tesla Philips Achieva MRI scanner with a 32-channel head coil. See Appendix 1 for more details.

Resting-state fMRI scans were co-registered with MPRAGE scans into the same anatomical space (native space); then 78 of the ROIs were parcellated on the rsfMRI scans, according to a multi-atlas fusion label algorithm (MALF) and large deformation diffeomorphic metric mapping, LDDMM [33,34]. Average time courses for the voxels in each ROI were normalized, and correlations between ROI pairs were calculated and normalized with the Fisher z-transformation. Of the 78 ROIs, we chose the ones that comprise the language-network ROIs and are functionally or structurally connected to the left IFG, the stimulated areaz0. In total 13 ROIs from the left hemisphere were selected. In addition, we conducted sensitivity analysis with an extended network that also includes the right homologues.

### 2.6 Statistical analyses

#### Prediction of potentially heterogeneous tDCS effects

In our prediction analysis, we modeled the additional, individual-specific tDCS effect using the so called modified outcome method, which adheres to established guidelines that account for heterogeneity in treatment effects [35–39]. For each participant, the individual-specific additional tDCS effect was defined as the difference between the potential change in the outcome if the participant had been assigned to active tDCS vs. sham tDCS, Y_i_^T = 1^ - Y_i_^T = 0^. We modeled the conditional average treatment effects (CATE), E[Y_i_^T = 1^ - Y_i_^T = 0^ | X_i_] = E[Y|T = 1, X] - E[Y|T = 0, X], via direct prediction modeling of a transformation U_i_ = 2Y_i_T_i_ – 2Y_i_(1 - T_i_) given the baseline covariates, X. This method replaced unobserved individual-specific additional tDCS effects, Y_i_^T = 1^ - Y_i_^T = 0^, with a fully observed modified outcome, U_i_, which allowed for prediction analysis of U given X. In addition, compared with the modeling analysis of E[Y|T, X], the modeling of E[U|X] avoided interaction terms between the treatment and covariates, thereby allowed valid prediction modeling with lower dimension, which was crucial given the modest sample size.

We conducted stepwise predictor selection based on cross-validation for the prediction of the modified outcome (see details in Appendix 2). Baseline covariates, i.e., the candidate predictors, were divided into multiple groups for further comparison across prediction models: 1) Demographic and clinical factors, including baseline semantic fluency, PPA variant, number of treatment sessions, sex, age, years post onset of symptoms, and total FTD-CDR severity and language severity measures. 2) Imaging factors, which consisted of correlations between the hypothesis-selected 13 language ROIs of the baseline rsfMRI; thus 78 ROI pairs (13 choose 2) in total. For sensitivity analysis, we also tested the functional and volumetric imaging factors within an extended network that involves right hemisphere areas as well (26 ROIs and thus 325 ROI pairs). The predictive R-squared (R^2^) and the root mean squared error (RMSE) are reported. Linear coefficients in the final prediction model are reported in Appendix 3 for diagnostic purposes.

## 3. Results

### 3.1 Prediction of potentially heterogeneous tDCS effects: clinical and demographic predictors

We tested the following non-imaging factors for predicting the additional tDCS effect on semantic fluency: baseline semantic fluency, PPA variant, number of treatment sessions, sex, age, years post onset of symptoms, and dementia and language severity (FTD-CDR overall sum and language measure, respectively) [29]. None of the non-imaging factors predicted the individual tDCS effect with R-squared increment thresholded at 0.1 (10%). Only dementia severity (overall FTD-CDR sum) marginally predicted the additional tDCS effect in semantic fluency with 4.4% increase in predictive R-squared, although RMSE did not improve after adding dementia severity into the model (Table 2). Even if one allows an additional round, the results remains similar: having lvPPA (or nfvPPA) provides only an additional 8.6% (or 5.4%) R2 increase, which leads to 13.0% (or 9.8%) accumulated R2 and RMSE 8.021 (or 8.166) with both overall FTD-CDR and having lvPPA (or nfvPPA) in the model. Therefore, neither lvPPA or nfvPPA would be selected in either the first or the second round. This result shows that PPA variant and severity of cognitive/language impairment cannot accurately predict individual tDCS effects.

**Table 2.** Non-imaging factors for individual tDCS effect prediction.

| | Accumulated $R^2$ | $R^2$ increase | RMSE |
| --- | --- | --- | --- |
| Null Model | 0 | 0 | 8.360 |
| Overall FTD-CDR | 0.044 | 0.044 | 8.407 |

### 3.2 Prediction of potentially heterogeneous tDCS effects: functional connectivity predictors

Since we did not find any demographic or clinical factor to predict the additional tDCS effect above the R-squared minimum threshold of 10%, we did not enter any other factors in the regression model to save degrees of freedom However, key baseline covariates via the literature and previous studies were included for completeness, including behavioral baseline performance.

The comparison between the final models from the non-imaging and imaging predictors (Tables 3 and 4) in terms of the accumulated predictive R^2^ (0.044 vs 0.498) and the RMSE (8.407 vs 6.287) indicated that the imaging predictors outweighed the non-imaging ones in predictive value.

**Table 3.** Imaging factors from the language network in the left hemisphere that predicted the individual tDCS effect. Predictiveness was evaluated by the LOOCV (predictive) R^2^.

| | Accumulated $R^2$ | $R^2$ increase | RMSE |
| --- | --- | --- | --- |
| Null Model | 0 | 0 | 8.496 |
| Left STG: Left MTG pole | 0.307 | 0.307 | 7.389 |
| Left IFG opercularis: Left IFG triangularis | 0.416 | 0.109 | 6.782 |
| Left IFG triangularis: Left AG | 0.498 | 0.082 | 6.287 |

**Table 4.** Imaging factors from the language network in both hemispheres that predicted the individual tDCS effect. Predictiveness was evaluated by the LOOCV (predictive) R^2^.

| | Accumulated $R^2$ | $R^2$ increase | RMSE |
| --- | --- | --- | --- |
| Null Model | 0 | 0 | 8.496 |
| Right AG : Right MTG | 0.380 | 0.380 | 6.989 |
| Left IFG triangularis : Left AG | 0.526 | 0.146 | 6.112 |
| Left IFG opercularis : Left IFG triangularis | 0.621 | 0.095 | 5.463 |

Consider specifically the left hemisphere, given that PPA is considered a LH syndrome. Two baseline resting-state functional connectivity ROI pairs predicted the additional tDCS effect on semantic fluency above 10% of R-squared increase (0.1 threshold): (1) the left superior temporal gyrus (STG)-to-left medial temporal gyrus (MTG) pole, and (2) left IFG opercularis-to-left IFG triangularis (Table 4 & Figure 2). Variable selection stopped at the last round with left IFG triangularis-to-left angular gyrus (AG) and just below 10% R-squared increase. The cumulative predictive R-squared of the final model was 49.8% (RMSE=6.29). The associated coefficients of the final model are reported in the supplementary materials (Appendix 3). Higher baseline connectivity in the first two pairs (most predictive, above 10% R^2^ increase) was associated with higher additional tDCS effect in the final model, whereas the trend was reversed in the last pair (8.2% R^2^ increase). In addition, we monitored all the pairs with above 10% R^2^ increases in each round of variable selection (see Figure 3).

**Figure 2.**
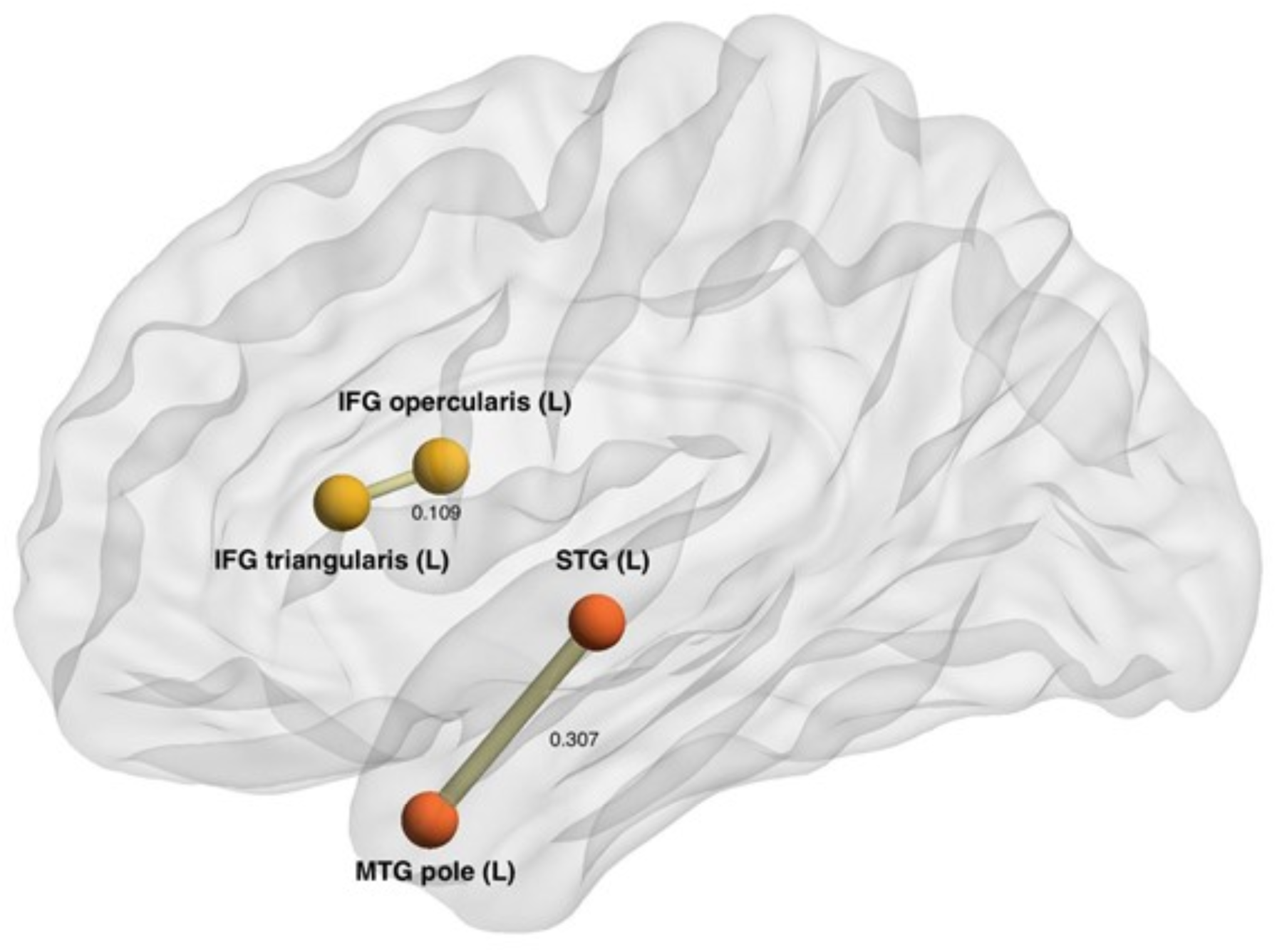
Visualization of the selected predictive imaging pairs for the additional tDCS effects. The positions of the nodes represent the average centers of each ROI from the cohort, rather than actual spatial distance. ROI pairs are plotted and connected if predictiveness of the baseline connectivity is confirmed by providing >10% R^2^ increase and being selected in the stepwise procedure. Thickness of the edge represents contribution to the accumulated R^2^ in the final prediction model.

**Figure 3.**
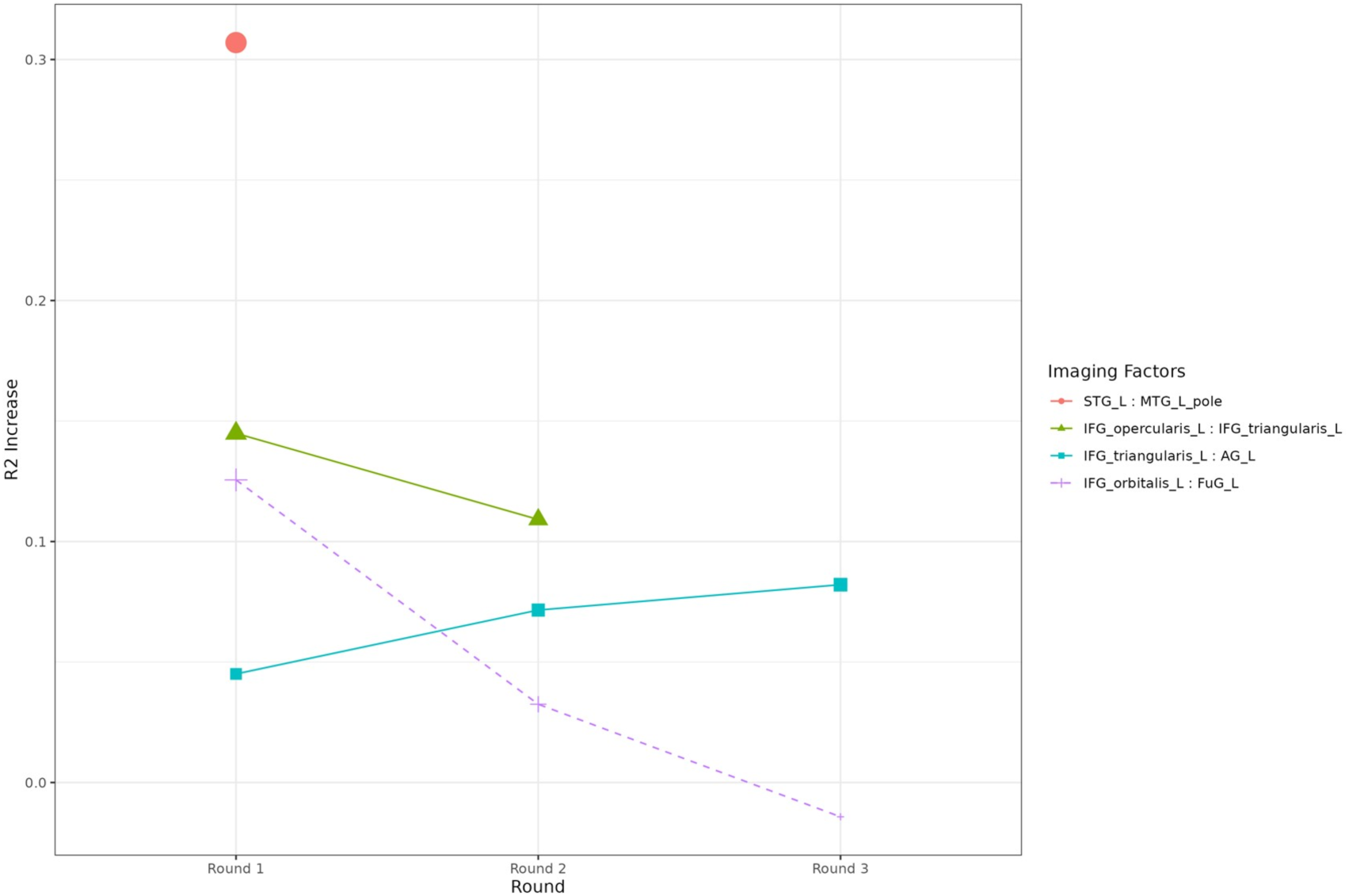
Functional connectivity pairs predicting the individual tDCS effects. Solid lines and points represent the selected factors in each round of the regression, whereas dotted lines represent the factors that were not selected but also provided over 0.1 increase of predictive R-squared in the first round. The 3 out of 4 pairs are the same as in Table 4. Note that in the first round another imaging predictor, Left IFG orbitalis: Left FuG provided R-squared increases greater than 0.1, but they were not selected because the Left STG: Left MTG pole had been selected for providing a larger R-squared increase.

In the sensitivity analysis that predicts individual treatment effects using the expanded connectivity network of 325 ROI pairs in both the LH and RH, three pairs were selected in each round, two overlapping with the main analysis (right AG-to-right MTG, left IFG triangularis-to-left AG, and left STG-to-left MTG pole, each providing R^2^ increases of 38.0%, 14.6%, and 9.5%; see Table 4 & Supplementary Table 3). The final prediction model showed improved prediction performance (RMSE 5.463, accumulated R^2^ 62.1%) compared to the LH-only prediction (RMSE 6.287, accumulated R^2^ 49.8%), as expected. In comparison, the volumetric predictors from the expanded network of 26 ROIs provided an accumulated R^2^ less than 10% (Appendix 4).

## 4. Discussion

The present study used a modified outcome prediction method that accounted for heterogeneity in PPA, to evaluate baseline clinical, demographic, behavioral (language) and imaging (rsFC and volumetric) predictors of tDCS effect on semantic fluency. Using the modified outcome addressed the problem of heterogeneity encountered in assessing treatment effects in this scenario and we conjecture that it would be similarly useful in any other heterogeneous neurodegenerative disorder. It was found that: (a) all clinical, demographic and behavioral baseline performance predictors together, accounted for 10% of the tDCS effect; (b) baseline volumes of these areas accounted only for 10% of the effect; (c) rsFC from a 7-min rsfMRI task, between temporal areas (left MTG pole to left STG) and frontal areas (left IFG pars opercularis to pars triangularis) predicted 50% of the tDCS effect on semantic fluency, when considering the LH alone, and, when considering both hemispheres, prediction improved for a total 62% of the tDCS effect on semantic fluency.

As far as we know, this is the first study that predicted a neuromodulatory effect (tDCS) in a neurodegenerative disorder and compared - in the same cohort, with the same method - other predictors of tDCS effect: clinical, demographic behavioral, and imaging (volumetric and rsFC) predictors. The present study has important implications for treatment prediction, advocating for rsFC as a biomarker for probable tDCS efficacy in PPA, and possibly for other neurodegenerative disorders, such as AD or MCI, with heterogeneous populations. We emphasize this important role that rsFC may play as a biomarker for patient capacity for treatment efficacy, rather than its typical role as an estimate of disease sequelae.

### 4.1 Semantic processing

An important finding in our study is that the strength of functional connectivity between temporal areas (MTG and STG), and between frontal areas (IFG opercularis and triangularis), predicted the magnitude of tDCS effect on semantic fluency. Lesion studies have shown that the left temporal cortex stores information about semantic categories, and frontal areas are important in accessing this information [40]. We have previously shown that the left ITG is responsible for storage of lexical characteristics of nouns and verbs in PPA [41,42], as well as with semantic fluency [43]. Furthermore, atrophy in the anterior and inferior left temporal regions, as well as in frontal regions, was associated with semantic fluency deficits [44]. The present study showed that baseline functional connectivity within the neural substrates of semantic fluency (semantic storage and control areas) predicts tDCS effects on semantic fluency.

### 4.2 Atrophy and FC considerations

The results of the present study seem counterintuitive given the atrophy in frontal areas in nfvPPA, who showed the largest tDCS effect [4]. Nevertheless, we [20] and others [45] have not found any correlation between functional connectivity and atrophy. Furthermore, we and others have found that active tDCS <u>improves</u> lower baseline function of atrophied regions by <u>reducing</u> the abnormally elevated connectivity (hyperconnectivity), to improve the efficiency of language processing [17–19]. This hyperconnectivity may be due to compensatory or disease-progression reasons, as previously proposed for the hippocampus [26,27]. It follows that active tDCS over hyperconnected areas may be beneficial. Unsurprisingly, in our previous clinical trial, patients with nfvPPA with the largest amount of atrophy in left IFG showed the largest benefits from IFG stimulation [32]. Therefore, active tDCS may be beneficial on compromised tissue (atrophied but still viable) when other network or compensatory brain areas remain intact. Finally, the present findings that hyperconnectivity between two semantic areas in the RH predict the tDCS effect, highlight the importance of RH semantic network areas, and their potential as tDCS targets.

### 4.3 Structural connectivity considerations

In the present study, we found that two of the most significant functional connectivity pairs that predicted the tDCS effect on semantic fluency, (Left IFG triangularis – Left IFG opercularis and Left MTG pole – Left STG) correspond to the end points of structurally connected areas because they are at the edges of the extreme capsule fasciculus, i.e., a white-matter bundle connecting the left IFG triangularis to temporal areas as part of the ventral language stream [46]. Several studies have shown that the extreme capsule is important for semantic processing and comprehension [46–48]. This bundle is sometimes hard to detect and often is considered as part of the uncinate fasciculus, although the latter has a more medial trajectory. We recently identified the white-matter integrity of this bundle as a significant predictor of tDCS effects but not language therapy alone (sham condition) for trained words during written naming [14]. In that study the contribution of the structural connectivity to the tDCS effect was significant, although smaller than in the present study (predicting only 12% of variance in outcome).

### 4.4 Limitations and implications

Certain limitations of this work should be noted. The most important one is the small sample size (36 patients with PPA), which, although not small for a neuromodulation randomized control trial in a rare neurodegenerative disorder such as PPA, is relatively small for robust conclusions in a prediction study.

To circumvent the sample size limitation, we used methods such as conservative stepwise linear predictor selection that aimed to discover the most predictive variables based on R^2^ increases of cross-validation predictiveness measures, rather than the multiple testing of hundreds of variables where multiplicity would be a primary concern. The second limitation is our lack of power to perform an analysis by PPA variants and thus there is a need to replicate these results in bigger samples. The novel heterogeneity analysis has helped alleviate these issues and provide a tool for prediction analyses in heterogeneous disorders. Finally, we have not considered any cerebellar regions, which could be interesting for future investigation.

## 5. Conclusion

The present study provides evidence that functional connectivity between areas of semantic processing (storage, control and working memory) better predict generalization of tDCS effects in semantic fluency than any other clinical, demographic, or imaging (volumetric and structural connectivity) factors in PPA. Functional connectivity was also found to be a better predictor for the main outcome than clinical, demographic, and volumetric predictors as shown in Appendix 5. These results have significant clinical implications. For example, baseline functional connectivity can serve as biomarker for selecting patients who will benefit from tDCS. These results demonstrate that a 7-minute rsfMRI scan of baseline resting-state connectivity is adequate to reliably predict which patient with PPA will benefit from neuromodulatory tDCS treatment.

## Data Availability

Data may be available upon request to the senior author.

## Acknowledgements

We would like to thank our participants and referring physicians for their dedication and interest in our study. Funding: This work was supported by the National Institutes of Health (National Institute of Deafness and Communication Disorders through award R01 DC014475 and National Institute of Aging through awards R01 AG068881, R01 AG075111 and R01 AG075404) to KT. AH was supported by NIH (NIDCD) through awards R01 DC05375, R01 DC011317 and P50 DC014664.

## Appendix 1.

T1-weighted MPRAGE sequence acquisition was performed according to the following parameters: a scan time of 6 minutes (150 slices); isotropic 1-mm voxel size; flip angle of 8°; SENSE acceleration factor of 2; TR/TE = 8/3.7 milliseconds (ms). Resting-state fMRI acquisition was performed according to the following parameters: scan time of 9 minutes (210 time-point acquisitions); slice thickness of 3 mm; in-plane resolution of 3.3x3.3 mm2; flip angle of 75°; SENSE acceleration factor of 2; SPIR for fat suppression; TR/TE = 2500/30 ms.

MPRAGE images were preprocessed and segmented into 283 regions of interest (ROIs) using MRICloud, a multi-atlas based, automated image parcellation approach. Preprocessing used a multi-atlas fusion label algorithm (MALF) and large deformation diffeomorphic metric mapping (LDDMM) [1,2], a highly accurate diffeomorphic algorithm that minimizes effects of atrophy or local space deformations on mapping. All images were processed in native space. Volumes for each ROI were normalized by total intracerebral volume (brain tissue excluding myelencephalon and cerebrospinal fluid) to control for relative regional atrophy.

Resting-state fMRI scans were preprocessed using MRICloud and included standard routines from the SPM connectivity toolbox for coregistration, motion, and slice timing correction; physiological nuisance correction using CompCor [3]; and motion and intensity TR outlier rejection using “ART” (https://www.nitrc.org/projects/artifact_detect/). To correct for motion, ART detected “outlier” TRs (2 standard deviations for motion and 4 standard deviations for intensity), which were used in combination with the physiological nuisance matrix in the deconvolution regression for the remaining TRs.

## Appendix 2: Predictor selection criteria

Throughout this manuscript we focus on stepwise linear predictor selection based on leave-one-out cross-validated (LOOCV) predictive R-squared (R^2^) that handles the sample size of this data.

Specifically, cross-validation was used in order to avoid biased evaluation of model predictiveness, and the goals of such variable selection were: 1) to discover the most predictive linear predictors, and 2) to compare the predictiveness of final prediction models built from different sets of predictors (such as non-imaging versus imaging factors) following a same variable selection procedure.

At each step, we test the increase of R^2^ after adding each of the variables that have not been added to the linear model, and select the variable with the largest R^2^ increase. Stepwise selection stops if the largest possible R^2^ increase of the last step is below 10%. The selected predictors till the last round are reported. Less important potential predictors may also be predictive if they are not selected into the model but have a larger than 10% R^2^ increase at a previous step, which might be correlated with the selected predictors and are of interest for future investigation. The predictive R-squared (R^2^) and the root mean squared error (RMSE) of each step, as well as the increase in R^2^ compared to the previous step, are reported. Linear coefficients in the final prediction model are reported in Appendix 3 for diagnostic purposes. The final prediction models built from non-imaging factors and imaging factors are compared by predictive R-squared and RMSE.

### Analysis of non-imaging (demographic and clinical) predictors

We first investigated whether any of the non-imaging factors (baseline semantic fluency, PPA variant, number of treatment sessions, sex, age, years post onset of symptoms, and total FTD-CDR severity and language severity measures) was predictive of the individual tDCS effect (quantified by pseudovalues).

### Analysis of imaging predictors

In this analysis, we tested the imaging factors (correlations between the hypothesis-selected 13 language ROIs of the baseline rsfMRI; this left hemisphere connectivity network consists of 78 ROI pairs in total). To handle missingness in the baseline resting-state functional connectivity data, an inverse propensity score weighting (IPW) method was applied. Propensity scores were estimated using logistic regression with the imaging missingness and baseline covariates. The inverse propensity scores weighted linear regression was then used in stepwise selection. The selection criteria based on LOOCV predictive R-squared increase for predicting pseudovalues remained the same.

As sensitivity analysis, we also tested the imaging factors within an expanded network that involves right hemisphere areas as well (26 ROIs and 325 ROI pairs in total). The predictive pairs were then compared with the language network analysis. The predictiveness of the final prediction model was still compared with non-imaging factors and language network factors. But less important potential predictors (that were not selected into the model and only provided >10% R^2^ increase at a previous step) are not reported for the reasons that 1) such aggressive variable selection beyond the conservative stepwise procedure would be too unstable given the numbers of complete cases (24) and candidate predictors (325) even when leave-one-out cross-validation is implemented, and 2) the goal of this sensitivity analysis was to confirm the reliability of the implemented stepwise selection procedure.

## Appendix 3.

In this appendix, we report the linear coefficients of each final prediction models for diagnostic purposes (non-imaging factors, language network factors, or expanded network factors that involve both language and DMN). Note that coefficients may be over-estimated, as the estimation was based the same data that was used for variable selection, and the p-values do not serve the usual purpose of valid hypothesis testing due to the selection bias in the estimates [4,5]. However, we report these statistics from the final prediction model fitting in order to be fully transparent for this study and provide information as a pilot study for possible directions of the associations.

**Supplementary Table 1.** Linear coefficients of the final prediction model using non-imaging factors.

|  | Estimate | SE | *t*(34) | *P* |
| --- | --- | --- | --- | --- |
| Intercept | 4.27 | 2.58 | 1.66 | 0.107 |
| Overall FTD-CDR | -0.59 | 0.30 | -1.95 | 0.059 |

**Supplementary Table 2.** **Linear coefficients of the final prediction model using left hemisphere language network imaging factors.**

|  | Estimate | SE | *t*(20) | *P* |
| --- | --- | --- | --- | --- |
| Intercept | -6.58 | 4.62 | -1.42 | 0.170 |
| Left STG : Left MTG pole | 31.27 | 7.05 | 4.44 | 0.000 |
| Left IFG opercularis : Left IFG triangularis | 12.50 | 4.67 | 2.68 | 0.014 |
| Left IFG triangularis : Left AG | -12.74 | 5.74 | -2.22 | 0.038 |

**Supplementary Table 3.** **Linear coefficients of the final prediction model using extended network (involving both left and right hemisphere) imaging factors.**

|  | Estimate | SE | *t*(20) | *P* |
| --- | --- | --- | --- | --- |
| Intercept | -5.32 | 3.14 | -1.69 | 0.106 |
| Right AG : Right MTG | 19.65 | 4.65 | 4.23 | 0.000 |
| Left IFG triangularis : Left AG | -17.01 | 4.83 | -3.52 | 0.002 |
| Left IFG opercularis : Left IFG triangularis | 20.37 | 6.63 | 3.07 | 0.006 |

## Appendix 4.

In this appendix, we report the sensitivity analysis with volumetric data in the extended network. Volumes of the ROIs are normalized by the intracerebral volume (total brain volume minus myelencephalon and cerebrospinal fluid).

None of the 26 ROIs provided more than 10% R^2^ increase in the first round (Left STG: 5.90%, Left SMG: 5.22%, Right AG: 0.91%), and even if we select Left STG and then enforce additional rounds of variable selection, only two ROIs can provide increase in R^2^ (Left SMG in the second round: 1.21%, Left IFG triangularis: 2.04%). This results in a prediction model with an accumulated R^2^ of 5.89% (it stops at the first round according to the selection criteria; if it exhausts the three rounds and only stops when no ROI provides R^2^ increase, it reaches 9.14%). For diagnostic purposes, the linear coefficients of the largest predictive model (the result of three rounds of selection so long as the predictor provides positive R^2^ changes) are reported below.

**Supplementary Table 4.** **Linear coefficients of the exhaustive search prediction model using extended network (involving both left and right hemisphere) volumetric data.**

|  | Estimate | SE | *t*(23) | *P* |
| --- | --- | --- | --- | --- |
| Intercept | -54.34 | 20.79 | -2.61 | 0.02 |
| Left STG | 1385.17 | 884.90 | 1.57 | 0.13 |
| Left SMG | 2502.73 | 1382.96 | 1.81 | 0.08 |
| Left IFG triangularis | 2603.51 | 2139.06 | 1.22 | 0.24 |

## Appendix 5.

In this appendix, we report the same analysis using the main outcome for trained items (letter counts for spelling).

**Supplementary Table 5.** **Non-imaging predictors for trained item individual tDCS effects.**

| | Accumulated $R^2$ | $R^2$ increase | RMSE |
| --- | --- | --- | --- |
| Null Model | 0 | 0 | 43.393 |
| Logopenic variant | 0.154 | 0.154 | 41.075 |

**Supplementary Table 6.** **Imaging predictors from the language network in the left hemisphere for trained item individual tDCS effects.**

| | Accumulated $R^2$ | $R^2$ increase | RMSE |
| --- | --- | --- | --- |
| Null Model | 0 | 0 | 43.861 |
| Left IFG opercularis : Left STG pole | 0.218 | 0.218 | 40.466 |
| Left IFG opercularis : Left IFG triangularis | 0.525 | 0.307 | 31.543 |
| Left IFG orbitalis : Left SMG | 0.628 | 0.103 | 27.916 |
| Left IFG orbitalis : Left MTG pole | 0.667 | 0.039 | 26.414 |

**Supplementary Table 7.** **Imaging predictors from the language network in both hemispheres for trained item individual tDCS effects.**

|  | Accumulated R<sup>2</sup> | R<sup>2</sup> increase | RMSE |
| --- | --- | --- | --- |
| Null Model | 0 | 0 | 43.861 |
| Left IFG opercularis : Left STG pole | 0.218 | 0.218 | 40.466 |
| Left IFG opercularis : Left IFG triangularis | 0.525 | 0.307 | 31.543 |
| Right STG pole : Right MTG pole | 0.676 | 0.151 | 26.056 |
| Left IFG opercularis : Left FuG | 0.752 | 0.076 | 22.778 |

**Supplementary Table 8.**
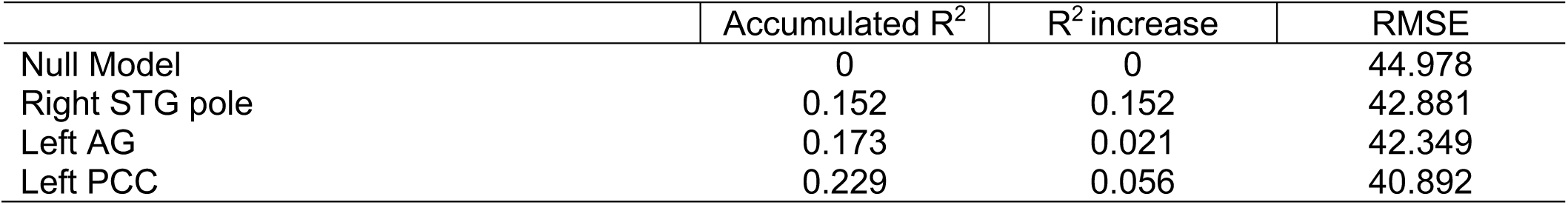
**Volumetric predictors after an exhaustive search from the language network in both hemispheres for trained item individual tDCS effects.**

**Supplementary Table 9.** **Linear coefficients of the final prediction model using non-imaging factors.**

|  | Estimate | SE | t(34) | P |
| --- | --- | --- | --- | --- |
| Intercept | 4.77 | 10.82 | 0.44 | 0.663 |
| Logopenic variant | -28.41 | 15.03 | -1.89 | 0.068 |

**Supplementary Table 10.** **Linear coefficients of the final prediction model using left hemisphere language network imaging factors.**

|  | Estimate | SE | t(20) | P |
| --- | --- | --- | --- | --- |
| Intercept | -41.34 | 15.02 | -2.75 | 0.013 |
| Left IFG opercularis : Left STG pole | -185.04 | 28.14 | -6.58 | 0.000 |
| Left IFG opercularis : Left IFG triangularis | 59.10 | 22.62 | 2.61 | 0.017 |
| Left IFG orbitalis : Left SMG | 81.58 | 24.55 | 3.32 | 0.004 |
| Left IFG orbitalis : Left MTG pole | 57.53 | 31.93 | 1.80 | 0.087 |

**Supplementary Table 11.**
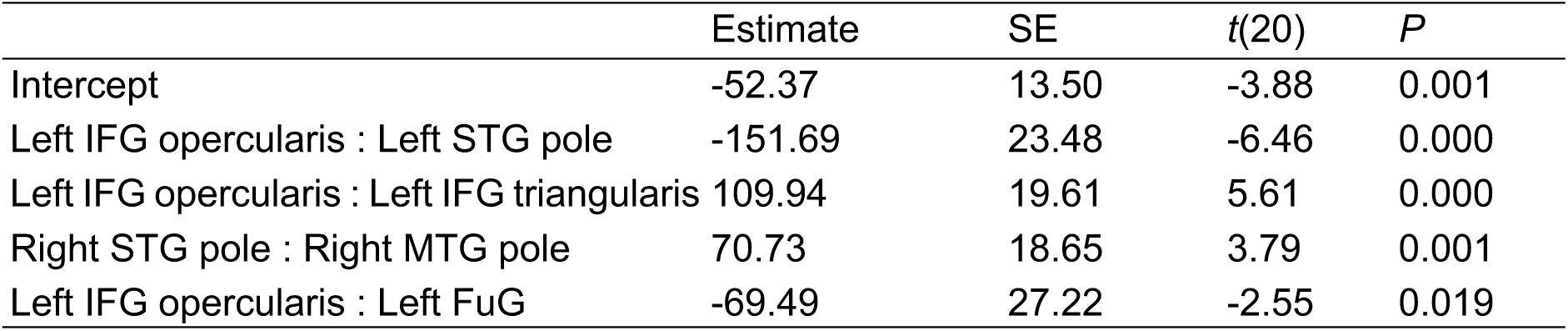
**Linear coefficients of the final prediction model using extended network (involving both left and right hemisphere) imaging factors.**

|  | Estimate | SE | *t*(20) | *P* |
| --- | --- | --- | --- | --- |
| Intercept | -52.37 | 13.50 | -3.88 | 0.001 |
| Left IFG opercularis : Left STG pole | -151.69 | 23.48 | -6.46 | 0.000 |
| Left IFG opercularis : Left IFG triangularis | 109.94 | 19.61 | 5.61 | 0.000 |
| Right STG pole : Right MTG pole | 70.73 | 18.65 | 3.79 | 0.001 |
| Left IFG opercularis : Left FuG | -69.49 | 27.22 | -2.55 | 0.019 |

**Supplementary Table 12.** **Linear coefficients of the exhaustive search prediction model using extended network (involving both left and right hemisphere) volumetric data.**

|  | Estimate | SE | *t*(23) | *P* |
| --- | --- | --- | --- | --- |
| Intercept | 136.64 | 76.74 | 1.78 | 0.092 |
| Right STG pole | -0.03 | 0.02 | -2.19 | 0.042 |
| Left AG | -0.03 | 0.01 | -2.07 | 0.053 |
| Left PCC | 0.01 | 0.01 | 1.78 | 0.092 |

## Notes

### Competing Interest Statement

The authors have declared no competing interest.

### Clinical Trial

NCT02606422

### Funding Statement

This study was funded by R01DC014475.

### Author Declarations

IRB of Johns Hopkins University School of Medicine gave ethical approval for this work (NA_00071337).

